# Age Based Trends for Unplanned Pediatric Rehospitalizations in the United States

**DOI:** 10.1101/2021.11.13.21266306

**Authors:** Nupur Amritphale, Amod Amritphale, Deepa Vasireddy, Mansi Batra, Mukul Sehgal, David Gremse

**Author notes:** Co-first authors: Contributed equally to all aspects of the research (Nupur Amritphale, Amod Amritphale).

## Abstract

**BACKGROUND AND OBJECTIVES:** Hospital readmission rate helps to highlight the effectiveness of post-discharge care. There remains a paucity of plausible age based categorization especially for ages below one year for hospital readmission rates.

**METHODS:** Data from 2017 Healthcare cost and utilization project National readmissions database was analyzed for ages 0-18 years. Logistic regression analysis was performed to identify predictors for unplanned early readmissions.

**RESULTS:** We identified 5,529,389 inpatient pediatric encounters which were further divided into age group cohorts. The overall rate of readmissions was identified at 3.2%. Beyond infancy, the readmission rate was found to be 6.7%. Across all age groups, the major predictors of unplanned readmission were cancers, diseases affecting transplant recipients and sickle cell patients. It was determined that reflux, milk protein allergy, hepatitis and inflammatory bowel diseases were significant comorbidities leading to readmission. Anxiety, depression and suicidal ideation depicted higher readmission rates in those older than 13 years. Across ages 1-4 yrs, dehydration, asthma and bronchitis were negative predictors of unplanned readmission.

**CONCLUSIONS:** Thirty-day unplanned readmissions remain a problem leading to billions of tax-payer-dollars lost per annum. Effective strategies for mandatory outpatient follow-up may help the financial aspect of care while also enhancing the quality of care.

## INTRODUCTION

Readmission rates have long been used by clinicians, hospital systems and health care commissions as a quality indicator. According to The Centers for Medicare and Medicaid services (CMS), about one in five Medicare patients discharged from the hospital were readmitted within 30 days. [1] With adequate measures and practicing preventive care, it has been seen that approximately 75% of them are potentially preventable. [2]

The readmission rates amongst adults have been vastly studied, however the pediatric world has a limited research to its name analyzing all causes. [3-4] Also it has been suggested that the pediatric data is becoming increasingly concentrated in large academic centers. [5] There is an increasing percentage of patients which are medically insured and there is also a better patient centered medical home provision along with accountable and coordinated care at multiple academic facilities. [6] Due to the improvement in neonatal-perinatal healthcare strategies, there is a vast majority of children who live longer with their chronic ailments and also develop long term consequences of the disease. This also however leads to an increase in the rate of unplanned readmissions. [7]

For this study, we used nationally representative data during the year 2017 to determine more recent trends in pediatric hospitalization and readmission. We have included all ages and stratified our study based on the age of patients admitted to determine age specific causes for readmissions. This study is composed of only unplanned readmissions and includes patients coming back within 30 days to give us a better picture of their etiology and therefore help us implement strategies in the future to limit readmissions.

## METHODS

The National Readmission Database (NRD) is a collection of all-age, all-payer discharges represented at the national level from U.S. non federal hospitals produced by the Healthcare Cost and Utilization Project of the Agency for Healthcare Research and Quality. [8] This database is composed of discharge-level hospitalization data from 28 geographically dispersed states across the United States of America. Approximately, it has over 5 million inpatient pediatric encounters for the year 2017 (weighted database for national representation). The dataset used in the present study represents 60% of the U.S. population and 58.2% of all U.S. hospitalizations. Every year, hospitalizations and rehospitalizations can be determined, using a de-identified unique patient linkage number assigned to each patient, which enables tracking of patients across hospitals within a state. Individual patients in the NRD are assigned up-to 40 diagnosis codes and 25 procedure codes for each hospitalization. The study was approved for an exempt status by the Institutional Review Board.

ICD-10 procedure & diagnosis codes (International Classification of Diseases, hereafter termed ICD-10 codes) and Clinical Classification Software refined (CCSR) codes were used to identify the pertinent discharge level diagnosis, procedures and organ system information. The data was divided into age groups 0 to 1 years; 1 to 4 years; 5 to 12 years and 13 to 18 years respectively for analysis purposes. The information for most common causes of admission for respective age groups was obtained from the Health cost and utilization project. Patients who died during their initial hospitalizations as well as those who were planned elective readmissions within 30 days of prior discharge were excluded. The cohort patients admitted in the month of December for the index admission were also excluded, as they may not have 30 days of follow-up, leading to immortal time bias.

ICD-10 codes and CCSR codes were used to define organ system disorders and various comorbidities. The All Patient Refined-Diagnosis Related Group (APR DRG) Mortality risk score and APR DRG Severity of Illness calculations were also performed in the analysis. The costs were determined by multiplying the hospital charges with the Agency for Healthcare Research and Quality’s all-payer cost-to-charge ratios for each hospital. All ICD-10 and CCSR codes used in this study shall be made available upon request. The causes of readmission were determined by the first diagnosis on the basis of Clinical Classification Software codes (CCSR).

Statistical analysis was performed using IBM SPSS Statistics for Windows, version 1.0.0.1327 (IBM Corp., Armonk, N.Y., USA) using 2-sided tests and a significance level of 0.05. Within the time periods, baseline characteristics of participants were examined and tested for statistical differences using the Pearson Chi Square test for categorical variables and Mann-Whitney U-Test for continuous variables with no readmission as the reference group. Logistic regressions were used to evaluate the predictors of readmission within respective age groups. The odds ratio was calculated by using logistic regression. We followed the methodology as has been previously described in well-written studies. [4, 9-14]

## RESULTS

### Population Characteristics and Descriptive Results

Our nationally registered database had a sample size of a total 5,529,389 inpatient encounters from January through December 2017 consisting of all ages from birth upto 18 years of age. The patients who died during their index admission as well as those who were discharged after November 30th were excluded from the index hospitalizations to remain within the threshold of 30-day readmissions. The remaining sample included a total of 5,096,320 patients who were discharged home alive. The total readmission rate for all age groups, all causes, was 3.2%. Readmission rates among infants were 2.1% while it tripled to 6.7% beyond infancy.

### Cost analysis and length of stay results

For ages below 1, the median hospital cost of index admission encounters were $1,363(IQR $894-$2,370) while for readmissions were $11,834 [IQR $5,738 - $23,471]. The median cost of index hospitalization for those with 30-day readmission were $1,734(IQR $1,734-$4,961) compared with $1,356(IQR $892-$2,343) for those with no readmission (P<0.001). The total charges for the index encounters and 30 days-unplanned readmission encounters amount to over $71 billion and $1.8 billion respectively. Median length of stay for index admission with and without 30 days readmission was 2 days (IQR 2-3 days) and 2 days (IQR 1-4 days) respectively. There was significant variation in cost and length of stay among different age groups (Table 1a), (Table 1b). Table 2 presents most frequent diagnosis associated with readmissions for all age groups.

**Table 1(a):** Baseline characteristics, demographics, organ system involvement and most common diagnosis for index pediatric admissions in the United States for age groups <1 and 1-4 years.

|  | Age <1 |  |  |  |  | Age 1-4 |  |  |  |  |
| --- | --- | --- | --- | --- | --- | --- | --- | --- | --- | --- |
|  | No early readmissions<br>(n= 3705889<br>; 97.9%) | 30 day readmission<br>(n= 75778<br>; 2.1%) | Overall<br>(n= 3781667) | P value | Odds ratio<br>(95% CI) | No early readmissions<br>(n= 264667<br>; 90.7%) | 30 day readmission<br>(n= 21230<br>; 9.3%) | Overall<br>(n= 285897) | P value | Odds ratio<br>(95% CI) |
| **Age(yrs) (median[IQR])** | 0 (0-0) | 0 (0-0) | 0 (0-0) | - |  | 2 (1-3) | 2 (1-3) | 2 (1-3) | - |  |
| Female | 1791312 (48.3%) | 32674 (43.1%) | 1823986 (48.2%) | <0.001 | 0.810<br>(0.799 - 0.822) | 114909 (43.4%) | 9203 (43.3%) | 124112 (43.4%) | 0.849 | 0.997<br>(0.970 - 1.026) |
| Elective | 56542 (1.5%) | 2615 (3.5%) | 59157 (1.6%) | <0.001 | 2.308<br>(2.218 - 2.402) | 46160 (17.5%) | 4283 (20.4%) | 50443 (17.7%) | <0.001 | 1.203<br>(1.162 - 1.246) |
| Weekend admission | 783985 (21.2%) | 17074 (22.5%) | 801059 (21.2%) | <0.001 | 1.084<br>(1.065 - 1.103) | 59725 (22.6%) | 4167 (19.6%) | 63892 (22.3%) | <0.001 | 0.838<br>(0.809 - 0.868) |
| **Primary expected payer** |  |  |  | <0.001 |  |  |  |  | <0.001 |  |
| Medicare | 15311 (0.4%) | 208 (0.3%) | 15519 (0.4%) |  |  | 898 (0.3%) | 156 (0.7%) | 1054 (0.4%) |  |  |
| Medicaid | 1759706 (47.5%) | 40726 (53.8%) | 1800432 (47.7%) |  |  | 162122 (61.4%) | 13033 (61.4%) | 175155 (61.4%) |  |  |
| Private | 1688188 (45.6%) | 30860 (40.8%) | 1719048 (45.5%) |  |  | 88449 (33.5%) | 7226 (34.1%) | 95675 (33.5%) |  |  |
| Self-pay | 136226 (3.7%) | 2002 (2.6%) | 138228 (3.7%) |  |  | 5415 (2%) | 295 (1.4%) | 5710 (2%) |  |  |
| No charge | 2446 (0.1%) | 27 (0%) | 2473 (0.1%) |  |  | 140 (0.1%) | 11 (0.1%) | 151 (0.1%) |  |  |
| Other | 100662 (2.7%) | 1890 (2.5%) | 102552 (2.7%) |  |  | 7193 (2.7%) | 491 (2.3%) | 7684 (2.7%) |  |  |
| **Quartile of median household income** |  |  |  | <0.001 |  |  |  |  | <0.001 |  |
| 0–25<sup>th</sup> | 881812 (24%) | 17716 (23.5%) | 899528 (23.9%) |  |  | 86453 (33%) | 6663 (31.6%) | 93116 (32.9%) |  |  |
| 26<sup>th</sup>–50<sup>th</sup> | 974100 (26.5%) | 20239 (26.9%) | 994339 (26.5%) |  |  | 73563 (28%) | 6292 (29.9%) | 79855 (28.2%) |  |  |
| 51<sup>st</sup>–75<sup>th</sup> | 1032625 (28.1%) | 21681 (28.8%) | 1054306 (28.1%) |  |  | 61114 (23.3%) | 5215 (24.8%) | 66329 (23.4%) |  |  |
| 76<sup>th</sup> –100<sup>th</sup> | 792419 (21.5%) | 15667 (20.8%) | 808086 (21.5%) |  |  | 41201 (15.7%) | 2894 (13.7%) | 44095 (15.6%) |  |  |
| **Comorbidities** |  |  |  |  |  |  |  |  |  |  |
| Dehydration | 53026 (1.4%) | 2660 (3.5%) | 55686 (1.5%) | <0.001 | 2.506<br>(2.409 -<br>2.607) | 50014 (18.9%) | 2873 (13.5%) | 52887 (18.5%) | <0.001 | 0.672<br>(0.645 -<br>0.699) |
| Cancer | 30864 (0.8%) | 1697 (2.2%) | 32561 (0.9%) | <0.001 | 2.728<br>(2.596 -<br>2.866) | 19304 (7.3%) | 7006 (33%) | 26310 (9.2%) | <0.001 | 6.261<br>(6.062 -<br>6.465) |
| Injury poisoning | 3806 (0.1%) | 50 (0.1%) | 3856 (0.1%) | 0.002 | 0.642<br>(0.486 -<br>0.849) | 2517 (1%) | 50 (0.2%) | 2567 (0.9%) | <0.001 | 0.246<br>(0.186 -<br>0.325) |
| Transplant | 310 (0%) | 50 (0.1%) | 360 (0%) | <0.001 | 7.892<br>(5.854 -<br>10.64)1 | 3242 (1.2%) | 923 (4.3%) | 4165 (1.5%) | <0.001 | 3.665<br>(3.402 -<br>3.949) |
| UTI | 15019 (0.4%) | 821 (1.1%) | 15840 (0.4%) | <0.001 | 2.692<br>(2.508 -<br>2.889) | 8657 (3.3%) | 787 (3.7%) | 9444 (3.3%) | 0.001 | 1.138<br>(1.057 -<br>1.226) |
| CKD_AKI_Nephritis | 6208 (0.2%) | 791 (1%) | 6999 (0.2%) | <0.001 | 6.286<br>(5.836 -<br>6.772) | 5523 (2.1%) | 949 (4.5%) | 6472 (2.3%) | <0.001 | 2.196<br>(2.046 -<br>2.356) |
| Sickle_cell | 4646 (0.1%) | 428 (0.6%) | 5074 (0.1%) | <0.001 | 4.525<br>(4.097 -<br>4.997) | 5879 (2.2%) | 526 (2.5%) | 6405 (2.2%) | 0.015 | 1.118<br>(1.022 -<br>1.224) |
| Cardiomyopathy_dysrhythmias | 11142 (0.3%) | 769 (1%) | 11911 (0.3%) | <0.001 | 3.400<br>(3.159 -<br>3.659) | 3137 (1.2%) | 500 (2.4%) | 3637 (1.3%) | <0.001 | 2.011<br>(1.828 -<br>2.212) |
| Intestinal_infection | 8794 (0.2%) | 606 (0.8%) | 9400 (0.2%) | <0.001 | 3.389<br>(3.120 -<br>3.681) | 14024 (5.3%) | 1122 (5.3%) | 15146 (5.3%) | 0.931 | 0.997<br>(0.937 -<br>1.062) |
| Appendiceal_conditions | –<sup>a</sup> | –<sup>a</sup> | –<sup>a</sup> | –<sup>a</sup> | –<sup>a</sup> | 2685 (1%) | 212 (1%) | 2897 (1%) | 0.824 | 0.984<br>(0.855 -<br>1.133) |
| IBD | –<sup>a</sup> | –<sup>a</sup> | –<sup>a</sup> | –<sup>a</sup> | –<sup>a</sup> | 258 (0.1%) | 23 (0.1%) | 281 (0.1%) | 0.627 | 1.111<br>(0.725 - |
|  |  |  |  |  |  |  |  |  |  | 1.703) |
| Hepatitis | _a | _a | _a | _a |  | 100 (0%) | 28 (0.1%) | 128 (0%) | <0.001 | 3.494<br>(2.297 -<br>5.314) |
| GI_other (digestive<br>hepto_biliary diseases) | 95814 (2.6%) | 7538 (9.9%) | 103352 (2.7%) | <0.001 | 4.162<br>(4.061 -<br>4.266) | 59392 (22.4%) | 8116 (38.2%) | 67508 (23.6%) | <0.001 | 2.139<br>(2.077 -<br>2.202) |
| Epilepsy | 13132 (0.4%) | 1581 (2.1%) | 14713 (0.4%) | <0.001 | 5.992<br>(5.684 -<br>6.316) | 30306 (11.5%) | 3630 (17.1%) | 33936 (11.9%) | <0.001 | 1.595<br>(1.536 -<br>1.656) |
| Diabetes | _a | _a | _a | _a | _a | 2224 (0.8%) | 98 (0.5%) | 2322 (0.8%) | <0.001 | 0.547<br>(0.447 -<br>0.670) |
| Endocrine_otherthan_diabetes | 92353 (2.5%) | 6936 (9.2%) | 99289 (2.6%) | <0.001 | 3.942<br>(3.843 -<br>4.044) | 78624 (29.7%) | 7647 (36%) | 86271 (30.2%) | <0.001 | 1.332<br>(1.294 -<br>1.372) |
| Cystic_fibrosis | _a | _a | _a | _a | _a | 999 (0.4%) | 51 (0.2%) | 1050 (0.4%) | 0.001 | 0.636<br>(0.480 -<br>0.842) |
| Skin_Subcutaneous | 53491 (1.4%) | 2589 (3.4%) | 56080 (1.5%) | <0.001 | 2.415<br>(2.320 -<br>2.514) | 36834 (13.9%) | 2638 (12.4%) | 39472 (13.8%) | <0.001 | 0.878<br>(0.841 -<br>0.915) |
| Psychiatric_disorders | 1090 (0%) | 96 (0.1%) | 1186 (0%) | <0.001 | 4.311<br>(3.499 -<br>5.312) | 2335 (0.9%) | 288 (1.4%) | 2623 (0.9%) | <0.001 | 1.545<br>(1.366 -<br>1.748) |
| Congenital_anomalies | 555947 (15%) | 18516 (24.4%) | 574463 (15.2%) | <0.001 | 1.832<br>(1.802 -<br>1.863) | 49195 (18.6%) | 5925 (27.9%) | 55120 (19.3%) | <0.001 | 1.696<br>(1.643 -<br>1.750) |
| Musculoskeletal | 7760 (0.2%) | 293 (0.4%) | 8053 (0.2%) | <0.001 | 1.850<br>(1.646 -<br>2.079) | 9046 (3.4%) | 444 (2.1%) | 9490 (3.3%) | <0.001 | 0.604<br>(0.548 -<br>0.665) |
| Asthma_bronchitis | 84371 (2.3%) | 4421 (5.8%) | 88792 (2.3%) | <0.001 | 2.659<br>(2.578 -<br>2.743) | 80663 (30.5%) | 4117 (19.4%) | 84780 (29.7%) | <0.001 | 0.549<br>(0.530 -<br>0.568) |
| Pneumonia | 23748 (0.6%) | 1535 (2%) | 25283 (0.7%) | <0.001 | 3.206<br>(3.043 -<br>3.377) | 36440 (13.8%) | 1919 (9%) | 38359 (13.4%) | <0.001 | 0.622<br>(0.593 -<br>0.653) |
| Respiratory_failure_arrest | 31998 (0.9%) | 3051 (4%) | 35049 (0.9%) | <0.001 | 4.817<br>(4.638 -<br>5.003) | 24218 (9.2%) | 2574 (12.1%) | 26792 (9.4%) | <0.001 | 1.370<br>(1.312 -<br>1.430) |
| Hemolytic_perinatal_jaundice | 824107 (22.2%) | 19913 (26.3%) | 844020 (22.3%) | <0.001 | 1.246<br>(1.226 -<br>1.267) | -<sup>a</sup> | -<sup>a</sup> | -<sup>a</sup> | -<sup>a</sup> | -<sup>a</sup> |
| Septicemia | 7841 (0.2%) | 606 (0.8%) | 8447 (0.2%) | <0.001 | 3.802<br>(3.499 -<br>4.131) | 4938 (1.9%) | 689 (3.2%) | 5627 (2%) | <0.001 | 1.764<br>(1.627 -<br>1.913) |
| Neutropenia | 9156 (0.2%) | 633 (0.8%) | 9789 (0.3%) | <0.001 | 3.401<br>(3.137 -<br>3.688) | 5140 (1.9%) | 1317 (6.2%) | 6457 (2.3%) | <0.001 | 3.339<br>(3.138 -<br>3.554) |
| **APR DRG Mortality risk** |  |  |  | <0.001 |  |  |  |  | <0.001 |  |
| Minor likelihood of dying | 3549349 (95.8%) | 65265 (86.1%) | 3614614 (95.6%) |  |  | 204279 (77.2%) | 10471 (49.3%) | 214750<br>(75.1%) |  |  |
| Moderate likelihood of dying | 97623 (2.6%) | 5510 (7.3%) | 103133 (2.7%) |  |  | 43188 (16.3%) | 7564 (35.6%) | 50752 (17.8%) |  |  |
| Major likelihood of dying | 42693 (1.2%) | 3704 (4.9%) | 46397 (1.2%) |  |  | 12792 (4.8%) | 2631 (12.4%) | 15423 (5.4%) |  |  |
| Extreme likelihood of dying | 11679 (0.3%) | 1197 (1.6%) | 12876 (0.3%) |  |  | 4000 (1.5%) | 552 (2.6%) | 4552 (1.6%) |  |  |
| **APR DRG Severity of Illness** |  |  |  | <0.001 |  |  |  |  | <0.001 |  |
| Minor loss of function (includes<br>cases with no comorbidity or<br>complications) | 2598278 (70.1%) | 42107 (55.6%) | 2640385 (69.8%) |  |  | 113472 (42.9%) | 3470 (16.3%) | 116942<br>(40.9%) |  |  |
| Moderate loss of function | 759309 (20.5%) | 18117 (23.9%) | 777426 (20.6%) |  |  | 90551 (34.2%) | 7388 (34.8%) | 97939 (34.3%) |  |  |
| Major loss of function | 297355 (8%) | 10736 (14.2%) | 308091 (8.1%) |  |  | 47172 (17.8%) | 7662 (36.1%) | 54834 (19.2%) |  |  |
| Extreme loss of function | 46402 (1.3%) | 4717 (6.2%) | 51119 (1.4%) |  |  | 13064 (4.9%) | 2699 (12.7%) | 15763 (5.5%) |  |  |
| **Hospital bed size** |  |  |  | <0.001 |  |  |  |  | <0.001 |  |
| Small | 595474 (16.1%) | 11315 (14.9%) | 606789 (16%) |  |  | 35944 (13.6%) | 3486 (16.4%) | 39430 (13.8%) |  |  |
| Medium | 1062727 (28.7%) | 17749 (23.4%) | 1080476 (28.6%) |  |  | 46616 (17.6%) | 2725 (12.8%) | 49341 (17.3%) |  |  |
| Large | 2047687 (55.3%) | 46714 (61.6%) | 2094401 (55.4%) |  |  | 182106 (68.8%) | 15019 (70.7%) | 197125<br>(68.9%) |  |  |
| **Control/Ownership of Hospital** |  |  |  | <0.001 |  |  |  |  | <0.001 |  |
| Government, nonfederal | 312803 (8.4%) | 8538 (11.3%) | 321341 (8.5%) |  |  | 31972 (12.1%) | 2488 (11.7%) | 34460 (12.1%) |  |  |
| Private, not-profit | 3123182 (84.3%) | 63317 (83.6%) | 3186499 (84.3%) |  |  | 217142 (82%) | 18190 (85.7%) | 235332 |  |  |
|  |  |  |  |  |  |  |  | (82.3%) |  |  |
| Private, invest-own | 269904 (7.3%) | 3923 (5.2%) | 273827 (7.2%) |  |  | 15553 (5.9%) | 552 (2.6%) | 16105 (5.6%) |  |  |
| **Hospital Urban Rural Designation** |  |  |  | <0.001 |  |  |  |  | <0.001 |  |
| Large metropolitan areas with at least 1 million residents | 1971291 (53.2%) | 43541 (57.5%) | 2014832 (53.3%) |  |  | 166418 (62.9%) | 14692 (69.2%) | 181110 (63.3%) |  |  |
| Small metropolitan areas with less than 1 million residents | 1407799 (38%) | 28025 (37%) | 1435824 (38%) |  |  | 86772 (32.8%) | 6236 (29.4%) | 93008 (32.5%) |  |  |
| Micropolitan areas | 255480 (6.9%) | 3385 (4.5%) | 258865 (6.8%) |  |  | 8322 (3.1%) | 235 (1.1%) | 8557 (3%) |  |  |
| Not metropolitan or micropolitan (non-urban residual) | 71319 (1.9%) | 826 (1.1%) | 72145 (1.9%) |  |  | 3155 (1.2%) | 66 (0.3%) | 3221 (1.1%) |  |  |
| **Hospital teaching status** |  |  |  | <0.001 |  |  |  |  | <0.001 |  |
| Metropolitan non-teaching hospital | 756340 (20.4%) | 14144 (18.7%) | 770484 (20.4%) |  |  | 19211 (7.3%) | 920 (4.3%) | 20131 (7%) |  |  |
| Metropolitan teaching hospital | 2622750 (70.8%) | 57422 (75.8%) | 2680172 (70.9%) |  |  | 233979 (88.4%) | 20009 (94.2%) | 253988 (88.8%) |  |  |
| Non-metropolitan hospital | 326799 (8.8%) | 4212 (5.6%) | 331011 (8.8%) |  |  | 11476 (4.3%) | 302 (1.4%) | 11778 (4.1%) |  |  |
| **Discharge destination** |  |  |  | <0.001 |  |  |  |  | <0.001 |  |
| Home/self-care | 3624806 (97.8%) | 71475 (94.3%) | 3696281 (97.7%) |  |  | 251799 (95.1%) | 18607 (87.7%) | 270406 (94.6%) |  |  |
| Home health care | 27262 (0.7%) | 1464 (1.9%) | 28726 (0.8%) |  |  | 2293 (0.9%) | 382 (1.8%) | 2675 (0.9%) |  |  |
| Discharge against medical advice | 887 (0%) | 29 (0%) | 916 (0%) |  |  | 169 (0.1%) | 15 (0.1%) | 184 (0.1%) |  |  |
| **Length of stay and cost analysis** |  |  |  |  |  |  |  |  |  |  |
| Index admission length of stay(days) (median[IQR]) | 2(2-3) | 2(2-3) | 2(2-3) |  |  | 2(1-4) | 3(2-6) | 2(1-4) |  |  |
| Index admission hospital charge (U.S.\$) (median[IQR]) | 4132(2724-7531) | 5419(3269-16106) | 4152(2732-7626) | | | 16194(8826-32798) | 27867(14161-60735) | 16814(9062-34493) | | |
| Index admission cost (U.S.\$) (median[IQR]) | 1356(892-2343) | 1734(1734-4961) | 1363(894-2370) | | | 4661(2626-9558) | 8373(4200-18828) | 4844(2691-10074) | | |
| Re-admission length of stay(days) (median[IQR]) |  |  | 2 [1 – 4] |  |  |  |  | 2 [2 – 5] |  |  |
| Re-admission hospital charge (U.S.\$) (median[IQR]) | | | 11834 [5738 – 23471] | | | | | 25072 [13494 – 49233] | | |
| Re-admission cost (U.S.\$) (median[IQR]) | | | 3820 [1934 – 7717] | | | | | 7583 [4019 – 14754] | | |

<sup>a</sup> : Per HCUP guidance, cell clusters with encounter numbers less than 10 were not reported

**Table 1(b):**
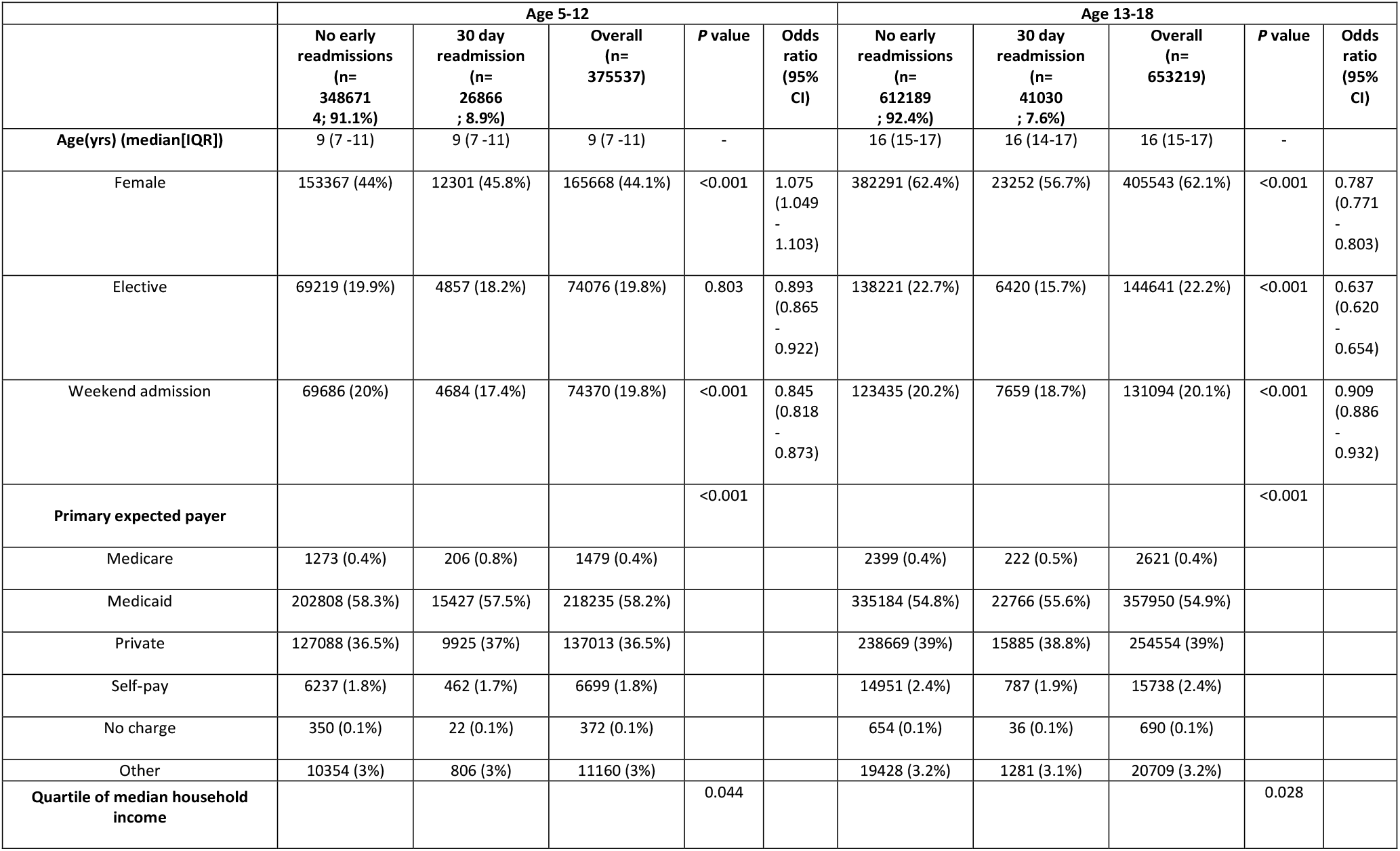
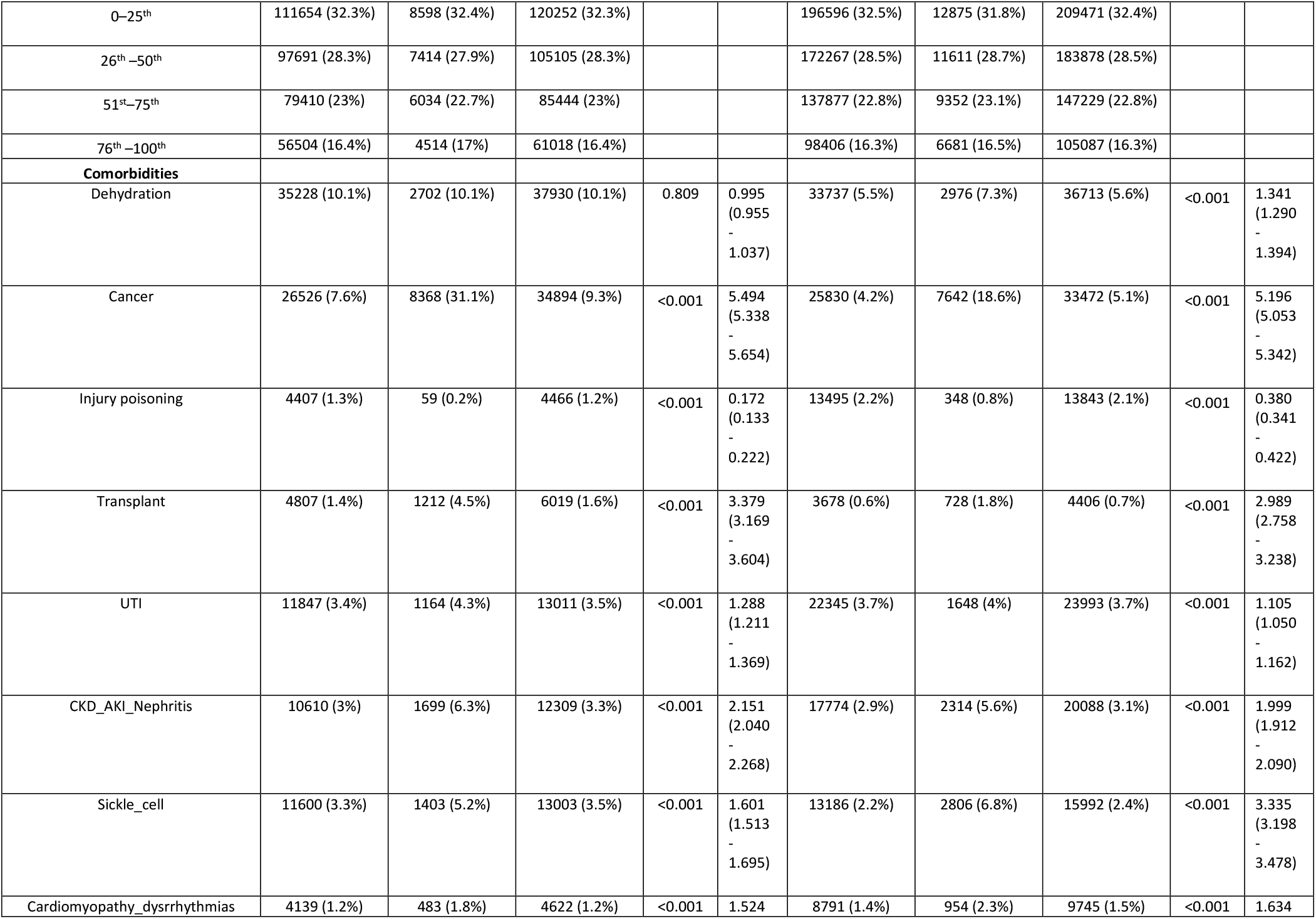
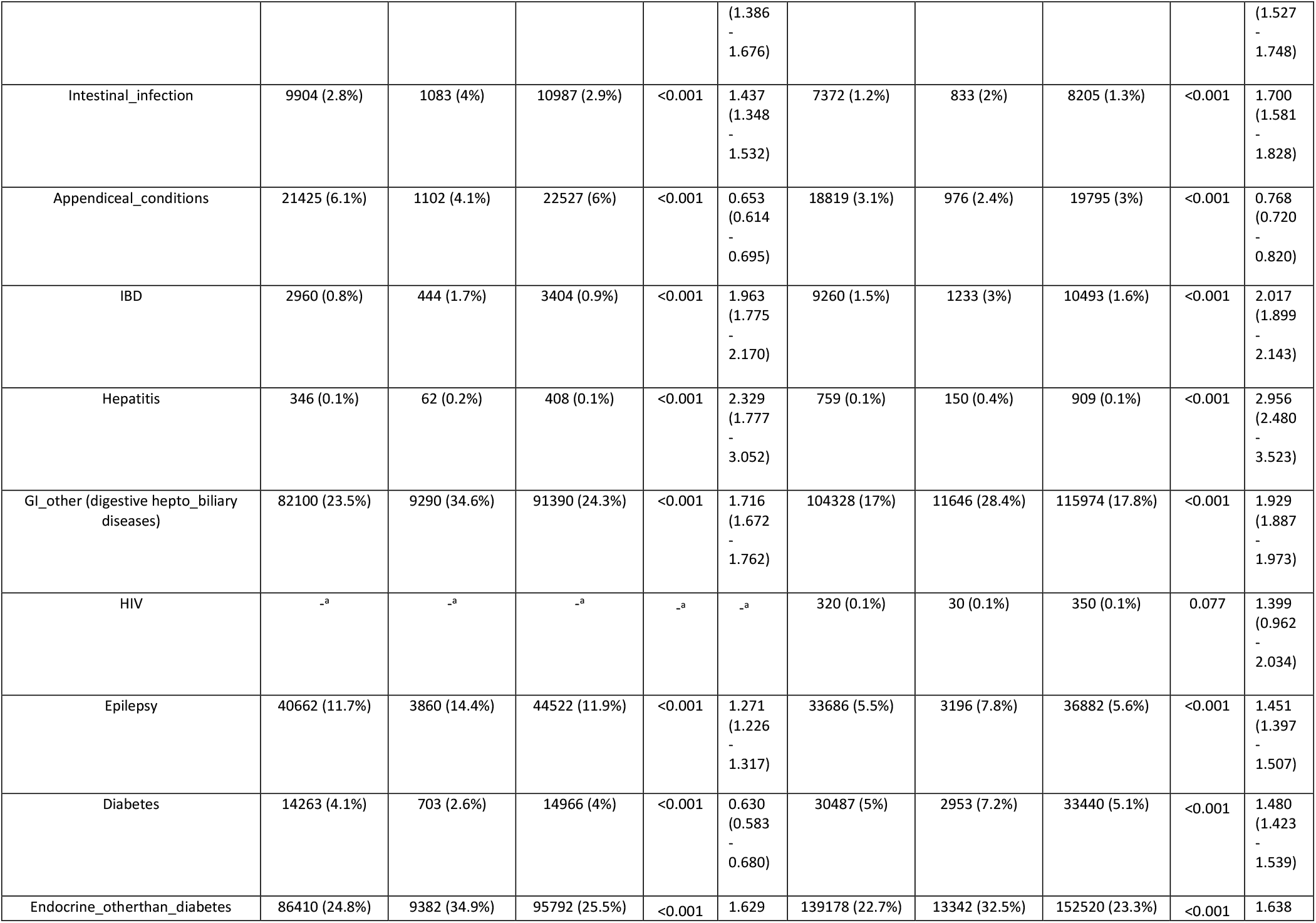
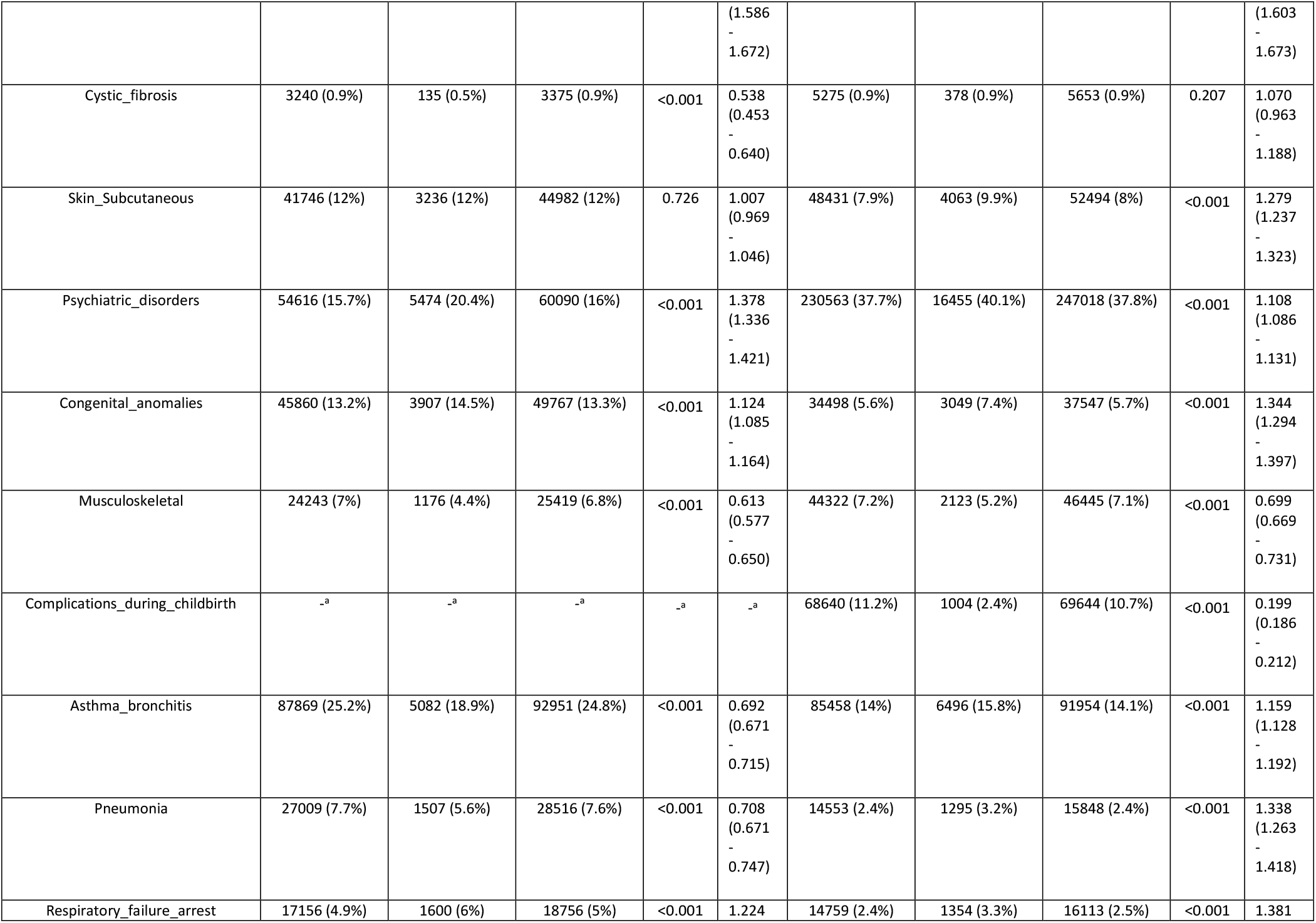
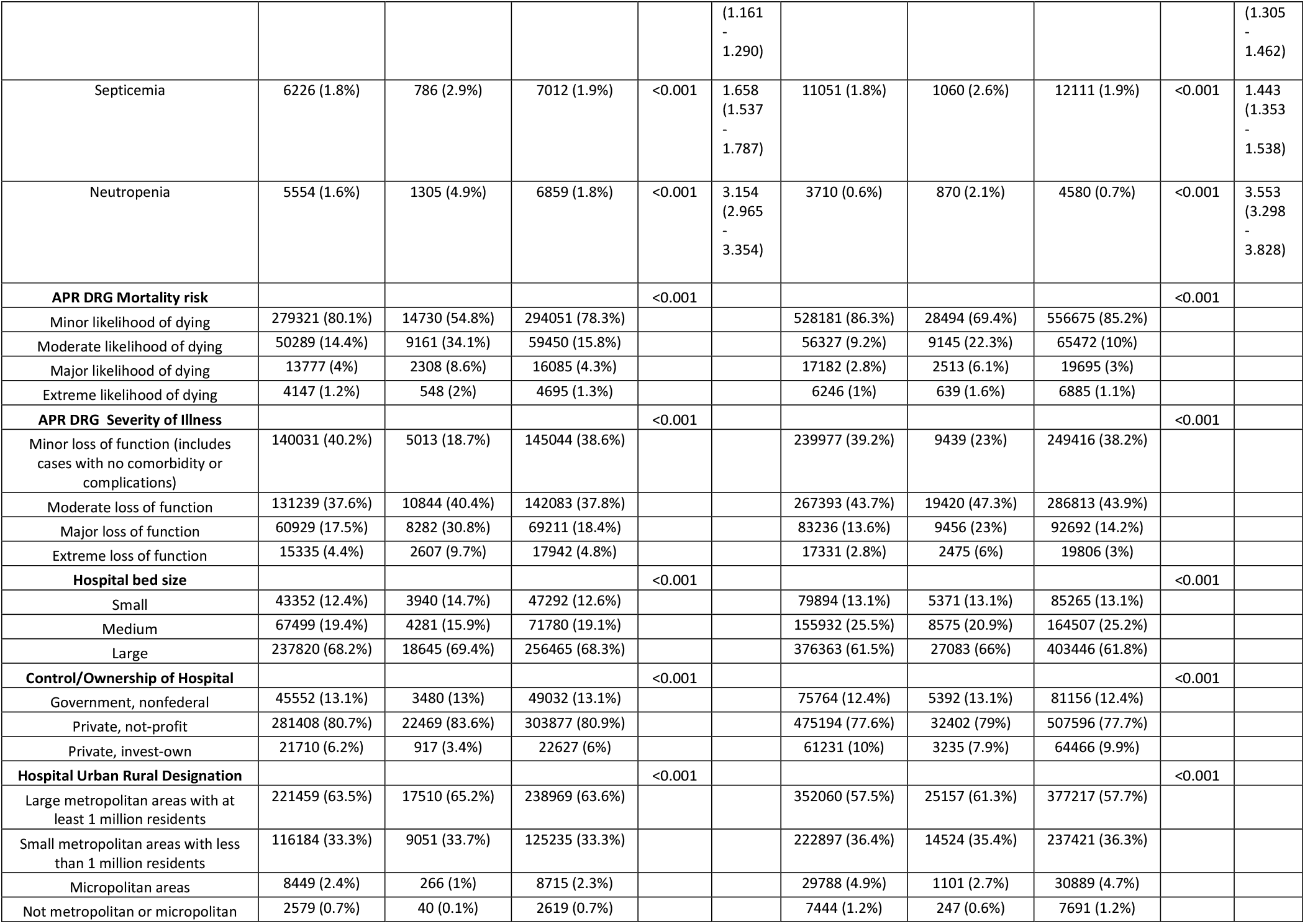
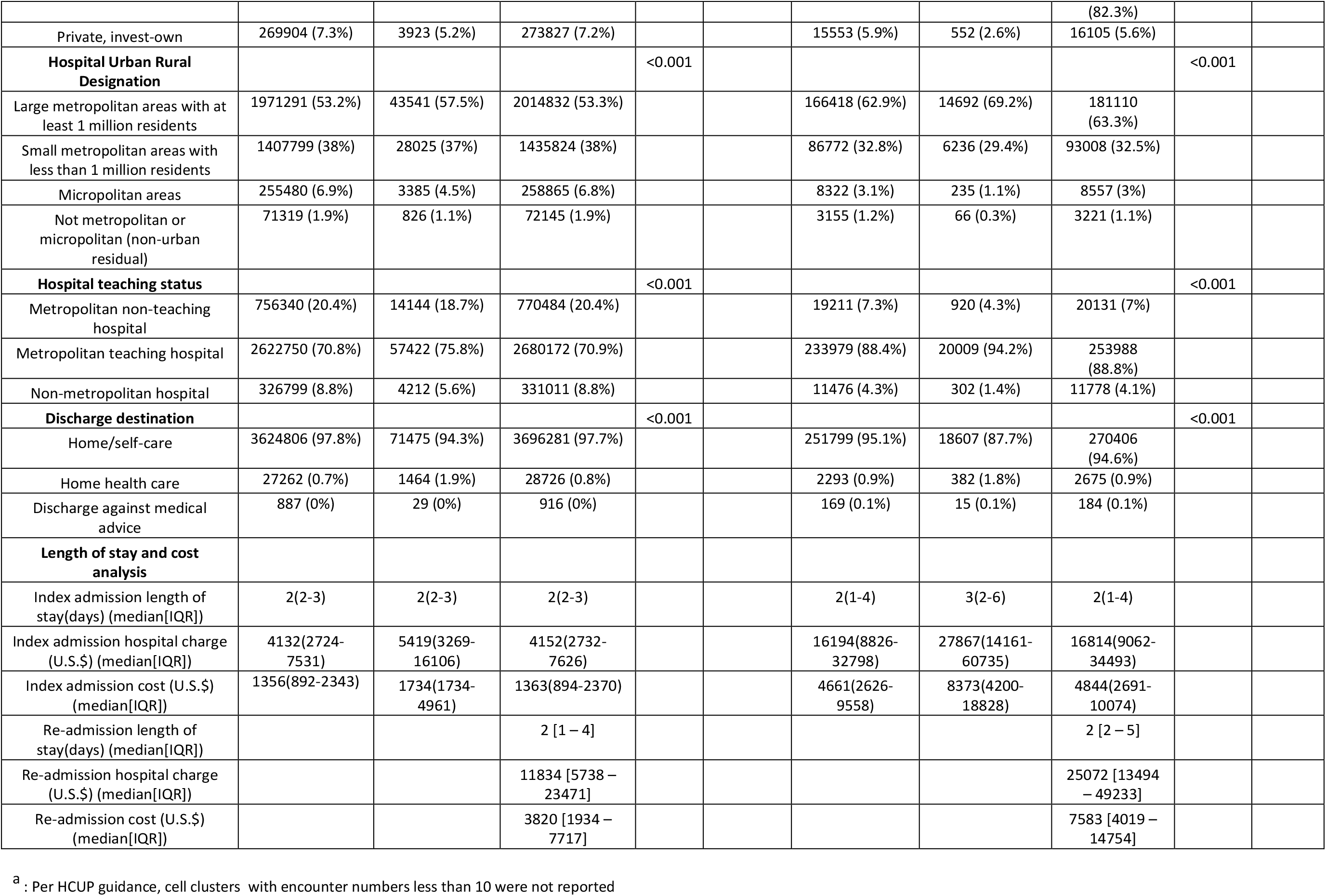
Baseline characteristics, demographics, organ system involvement and most common diagnosis for index pediatric admissions in the United States for age groups 5-12 years and 13-18 years.

**Table 2:** Causes and frequencies of primary diagnosis category for readmissions encounters. [Based on the primary Clinical Classification Software refined (CCSR) codes]

| Age <1 |  | Age 1-4 |  | Age 5-12 |  | Age 13-18 |  |
| --- | --- | --- | --- | --- | --- | --- | --- |
| Diagnosis category | Frequency (%)<br>n=76,274 | Diagnosis category | Frequency (%)<br>n=21,340 | Diagnosis category | Frequency (%)<br>n=26,723 | Diagnosis category | Frequency (%)<br>n=41,180 |
| Hemolytic jaundice | 20338 (26.7%) | Conditions due to neoplasm or the treatment of neoplasm/antineoplastic therapies | 3157 (14.8%) | Conditions due to neoplasm or the treatment of neoplasm antineoplastic therapies | 3995 (15%) | Depressive disorders | 4696 (11.4%) |
| Other unspecified perinatal conditions | 9140 (12%) | Diseases of white blood cells | 1390 (6.5%) | Complication of other surgical or medical care, injury, initial encounter | 1533 (5.7%) | Conditions due to neoplasm or the treatment of neoplasm / antineoplastic therapies | 4083 (9.9%) |
| Perinatal infections | 6960 (9.1%) | Complication of other surgical or medical care, injury, initial encounter | 1176 (5.5%) | Diseases of white blood cells | 1447 (5.4%) | Sickle cell trait/anemia | 2303 (5.6%) |
| Acute bronchitis | 5236 (6.9%) | Epilepsy; convulsions | 1022 (4.8%) | Other specified and unspecified mood disorders | 1019 (3.8%) | Bipolar and related disorders | 1753 (4.3%) |
| Neonatal digestive and feeding disorders | 3223 (4.2%) | Pneumonia (except that caused by tuberculosis) | 945 (4.4%) | Sickle cell trait/anemia | 1008 (3.8%) | Diabetes mellitus with complication | 1717 (4.2%) |
| Respiratory perinatal condition | 2614 (3.4%) | Respiratory failure; insufficiency; arrest | 932 (4.4%) | Epilepsy; convulsions | 884 (3.3%) | Complication of other surgical or medical care, injury, initial encounter | 1428 (3.5%) |
| Digestive congenital anomalies | 2075 (2.7%) | Acute bronchitis | 859 (4%) | Depressive disorders | 706 (2.6%) | Other specified and unspecified mood disorders | 1379 (3.3%) |
| Other general signs and symptoms | 1989 (2.6%) | Asthma | 852 (4%) | Pneumonia (except that caused by tuberculosis) | 695 (2.6%) | Diseases of white blood cells | 942 (2.3%) |
| Respiratory failure; insufficiency; arrest | 1709 (2.2%) | Other specified upper respiratory infections | 758 (3.6%) | Asthma | 617 (2.3%) | Schizophrenia spectrum and other psychotic disorders | 849 (2.1%) |
| Other specified upper respiratory infections | 1258 (1.6%) | Disorders of fluid and electrolytes | 745 (3.5%) | Intestinal infections | 533 (2%) | Complications during the puerperium | 772 (1.9%) |

### Predictors of 30-day readmission

Table 3 presents the results of logistic regression analysis for the predictors of 30-day readmission for all age groups. For children less than 1 year old, the overall comorbidities for pediatric readmission in this age group, excluding live birth, were epilepsy/seizures [2.707 (2.556 - 2.866), p<0.001], gastrointestinal ailments including reflux/milk protein allergy [2.264 (2.199 - 2.330), p<0.001] and sickle cell disease [2.625 (2.369 - 2.909), p<0.001]. Bronchiolitis [1.731 (1.667 - 1.798), p<0.001] and hemolytic anemia [1.306 (1.285 - 1.328), p<0.001] were also having increased risks of readmissions associated with this group. It was also seen that dehydration, septicemia, cystic fibrosis and injury/poisonings were negative predictors for readmission in this age group.

**Table 3:** Logistic regression analysis for 30 day readmissions.

|  | Age <1 |  | Age 1-4 |  | Age 5-12 |  | Age 13-18 |  |
| --- | --- | --- | --- | --- | --- | --- | --- | --- |
|  | *P*<br>value | Odds ratio | *P* value | Odds ratio | *P* value | Odds ratio | *P* value | Odds ratio |
| **Primary expected payer**<br>(Reference: Medicaid) |  |  |  |  |  |  |  |  |
| Medicare | <0.001 | 0.588 (0.512 - 0.674) | <0.001 | 2.166 (1.826 - 2.569) | <0.001 | 2.127 (1.834 - 2.467) | <0.001 | 1.364 (1.188 - 1.566) |
| Private | <0.001 | 0.790 (0.778 - 0.802) | 0.289 | 1.016 (0.986 - 1.047) | 0.049 | 1.027 (1.000 - 1.054) | 0.005 | 0.980 (0.960 - 1.001) |
| Self-pay | <0.001 | 0.635 (0.607 - 0.664) | <0.001 | 0.678 (0.602 - 0.763) | 0.575 | 0.973 (0.884 - 1.071) | <0.001 | 0.775 (0.720 - 0.834) |
| No charge | <0.001 | 0.475 (0.325 - 0.694) | 0.994 | 1.002 (0.546 - 1.839) | 0.340 | 0.809 (0.523 - 1.251) | 0.216 | 0.808 (0.577 - 1.131) |
| Other | <0.001 | 0.811 (0.774 - 0.850) | 0.001 | 0.850 (0.774 - 0.933) | 0.550 | 1.023 (0.950 - 1.101) | 0.300 | 0.971 (0.916 - 1.029) |
| **Quartile of median household income**<br>(Reference: 0-25<sup>th</sup> centile) |  |  |  |  |  |  |  |  |
| 26<sup>th</sup>–50<sup>th</sup> | 0.001 | 1.034 (1.013 - 1.055) | <0.001 | 1.110 (1.071 - 1.150) | 0.379 | 0.986 (0.954 - 1.018) | 0.003 | 1.029 (1.003 - 1.056) |
| 51<sup>st</sup>–75<sup>th</sup> | <0.001 | 1.045 (1.024 - 1.066) | <0.001 | 1.107 (1.066 - 1.150) | 0.444 | 0.987 (0.954 - 1.021) | 0.001 | 1.036 (1.008 - 1.065) |
| 76<sup>th</sup>–100<sup>th</sup> | 0.148 | 0.984 (0.963 - 1.006) | <0.001 | 0.911 (0.871 - 0.953) | 0.054 | 1.037 (0.999 - 1.077) | 0.001 | 1.037 (1.006 - 1.069) |
| **Comorbidities**<br>(Reference: no) |  |  |  |  |  |  |  |  |
| Dehydration | <0.001 | 0.794 (0.755 - 0.836) | <0.001 | 0.636 (0.603 - 0.670) | <0.001 | 0.807 (0.767 - 0.848) | <0.001 | 0.861 (0.823 - 0.900) |
| Cancer | <0.001 | 1.745 (1.657 - 1.836) | <0.001 | 6.217 (5.989 - 6.454) | <0.001 | 5.453 (5.278 - 5.634) | <0.001 | 4.892 (4.745 - 5.043) |
| Injury/Poisoning | <0.001 | 0.382 (0.279 - 0.522) | <0.001 | 0.476 (0.352 - 0.644) | <0.001 | 0.323 (0.248 - 0.421) | <0.001 | 0.589 (0.523 - 0.663) |
| Transplant |  | 2.061 (1.502 - 2.830) | <0.001 | 1.942 (1.789 - 2.108) | <0.001 | 1.704 (1.587 - 1.829) | <0.001 | 1.400 (1.284 - 1.526) |
| UTI | <0.001 | 1.307 (1.212 - 1.408) | <0.001 | 1.026 (0.949 - 1.110) | <0.001 | 1.190 (1.115 - 1.270) | 0.523 | 0.983 (0.932 - 1.037) |
| Renal disorders | <0.001 | 1.697 (1.563 - 1.843) | <0.001 | 1.553 (1.437 - 1.677) | <0.001 | 1.668 (1.572 - 1.770) | <0.001 | 1.451 (1.381 - 1.525) |
| Sickle cell disease | <0.001 | 2.625 (2.369 - 2.909) | <0.001 | 1.678 (1.530 - 1.841) | <0.001 | 2.413 (2.275 - 2.560) | <0.001 | 3.890 (3.721 - 4.066) |
| Cardiomyopathy and dysrhythmias | <0.001 | 1.339 (1.237 - 1.449) | <0.001 | 1.367 (1.234 - 1.515) | 0.007 | 1.149 (1.038 - 1.272) | <0.001 | 1.219 (1.136 - 1.309) |
| Intestinal infection | <0.001 | 1.416 (1.295 - 1.549) | 0.081 | 0.941 (0.878 - 1.008) | 0.013 | 1.093 (1.019 - 1.172) | 0.005 | 1.117 (1.035 - 1.206) |
| Appendiceal conditions | - | - | <0.001 | 1.332 (1.152 - 1.541) | 0.101 | 1.056 (0.990 - 1.126) | 0.277 | 0.964 (0.901 - 1.030) |
| IBD | - | - | 0.903 | 0.973 (0.623 - 1.518) | <0.001 | 2.131 (1.920 - 2.366) | <0.001 | 1.893 (1.778 - 2.017) |
| Hepatitis | - | - | <0.001 | 1.770 (1.096 - 2.859) | <0.001 | 1.702 (1.286 - 2.254) | <0.001 | 1.779 (1.483 - 2.135) |
| Digestive and hepatobiliary diseases | <0.001 | 2.264 (2.199 - 2.330) | <0.001 | 1.658 (1.606 - 1.712) | <0.001 | 1.417 (1.377 - 1.458) | <0.001 | 1.427 (1.393 - 1.463) |
| Epilepsy | <0.001 | 2.707 (2.556 - 2.866) | <0.001 | 1.622 (1.557 - 1.690) | <0.001 | 1.439 (1.384 - 1.497) | <0.001 | 1.339 (1.287 - 1.394) |
| Diabetes | 0.064 | 0.472 (0.214 - 1.044) | <0.001 | 0.717 (0.582 - 0.884) | <0.001 | 0.841 (0.776 - 0.910) | <0.001 | 1.468 (1.407 - 1.531) |
| Endocrine causes other than diabetes | <0.001 | 1.948 (1.878 - 2.020) | <0.001 | 1.360 (1.308 - 1.415) | <0.001 | 1.383 (1.339 - 1.429) | <0.001 | 1.246 (1.214 - 1.278) |
| Cystic fibrosis | 0.001 | 0.117 (0.056 - 0.246) | 0.024 | 0.721 (0.543 - 0.958) | <0.001 | 0.675 (0.566 - 0.804) | <0.001 | 0.832 (0.745 - 0.928) |
| Skin and Subcutaneous | <0.001 | 1.342 (1.286 - 1.400) | <0.001 | 0.898 (0.859 - 0.939) | <0.001 | 0.929 (0.893 - 0.967) | <0.001 | 0.961 (0.928 - 0.995) |
| Psychiatric disorders | <0.001 | 1.320 (1.063 - 1.640) | 0.705 | 0.975 (0.855 - 1.112) | <0.001 | 1.634 (1.580 - 1.689) | <0.001 | 1.272 (1.244 - 1.300) |
| Congenital anomalies | <0.001 | 1.449 (1.423 - 1.475) | <0.001 | 1.554 (1.501 - 1.610) | <0.001 | 1.108 (1.067 - 1.152) | <0.001 | 1.135 (1.090 - 1.182) |
| Musculoskeletal | 0.577 | 0.962 (0.839 - 1.103) | <0.001 | 0.736 (0.661 - 0.819) | <0.001 | 0.777 (0.728 - 0.828) | <0.001 | 0.779 (0.741 - 0.819) |
| Complications during childbirth |  | - | - | - | - | - | <0.001 | 0.294 (0.275 - 0.313) |
| Asthma and bronchitis | <0.001 | 1.731 (1.667 - 1.798) | <0.001 | 0.870 (0.837 - 0.904) | <0.001 | 0.881 (0.852 - 0.911) | 0.078 | 1.026 (0.997 - 1.056) |
| Pneumonia | 0.007 | 1.170 (1.102 - 1.241) | <0.001 | 0.790 (0.750 - 0.832) | <0.001 | 0.773 (0.730 - 0.819) | 0.520 | 0.979 (0.919 - 1.044) |
| Respiratory failure and arrest | <0.001 | 1.476 (1.406 - 1.550) | <0.001 | 1.398 (1.331 - 1.468) | <0.001 | 1.235 (1.163 - 1.310) | 0.003 | 1.101 (1.033 - 1.173) |
| Hemolytic and perinatal jaundice | <0.001 | 1.306 (1.285 - 1.328) | - | - | - | - | - | - |
| Septicemia | <0.001 | 0.802 (0.731 - 0.880) | 0.295 | 0.953 (0.872 - 1.043) | <0.001 | 0.936 (0.862 - 1.017) | 0.002 | 0.896 (0.835 - 0.961) |
| Neutropenia | <0.001 | 1.516 (1.393 - 1.650) | 0.830 | 1.008 (0.940 - 1.080) | <0.001 | 0.982 (0.918 - 1.050) | 0.218 | 1.052 (0.971 - 1.139) |
| **APR DRG Mortality risk**<br>(Reference: minor) |  |  |  |  |  |  |  |  |
| Moderate likelihood of dying | <0.001 | 3.069 (2.984 - 3.157) | <0.020 | 3.417 (3.312 - 3.526) | <0.001 | 3.454 (3.360 - 3.552) | <0.001 | 3.010 (2.935 - 3.086) |
| Major likelihood of dying | <0.001 | 4.719 (4.559 - 4.884) | <0.001 | 4.013 (3.831 - 4.203) | <0.001 | 3.177 (3.031 - 3.330) | <0.001 | 2.711 (2.596 - 2.832) |
| Extreme likelihood of dying | <0.001 | 5.574 (5.250 - 5.919) | <0.001 | 2.694 (2.459 - 2.951) | <0.001 | 2.505 (2.288 - 2.742) | <0.001 | 1.897 (1.748 - 2.060) |
| **APR DRG Severity of Illness**(Reference: minor) |  |  |  |  |  |  |  |  |
| Moderate loss of function | <0.001 | 1.472 (1.447 - 1.498) | <0.001 | 2.668 (2.560 - 2.780) | <0.001 | 2.308 (2.230 - 2.389) | <0.001 | 1.846 (1.800 - 1.894) |
| Major loss of function | <0.001 | 2.228 (2.180 - 2.276) | <0.001 | 5.311 (5.095 - 5.536) | <0.001 | 3.797 (3.662 - 3.938) | <0.001 | 2.888 (2.804 - 2.975) |
| Extreme loss of function | <0.001 | 6.272 (6.078 - 6.473) | <0.001 | 6.755 (6.403 - 7.126) | <0.001 | 4.749 (4.517 - 4.994) | <0.001 | 3.631 (3.465 - 3.805) |
| **Hospital bed size**<br>(Reference: small) |  |  |  |  |  |  |  |  |
| Medium | <0.001 | 0.879 (0.858 - 0.900) | <0.001 | 0.603 (0.572 - 0.635) | <0.001 | 0.698 (0.667 - 0.730) | <0.001 | 0.818 (0.790 - 0.847) |
| Large | <0.001 | 1.201 (1.176 - 1.226) | <0.001 | 0.850 (0.818 - 0.884) | <0.001 | 0.863 (0.832 - 0.894) | <0.001 | 1.070 (1.038 - 1.103) |
| **Control/Ownership of Hospital** (Reference: government, nonfederal) |  |  |  |  |  |  |  |  |
| Private, not-profit | <0.001 | 0.743 (0.726 - 0.760) | 0.001 | 1.077 (1.031 - 1.124) | 0.020 | 1.045 (1.007 - 1.085) | 0.001 | 0.958 (0.930 - 0.987) |
| Private, invest-own | <0.001 | 0.532 (0.513 - 0.553) | <0.001 | 0.456 (0.415 - 0.502) | <0.001 | 0.553 (0.513 - 0.596) | <0.001 | 0.742 (0.710 - 0.776) |
| **Hospital Urban Rural Designation**<br>(Reference:Large metropolitan areas with at least 1 million residents) |  |  |  |  |  |  |  |  |
| Small metropolitan areas with less than 1 million residents | <0.001 | 0.901 (0.888 - 0.915) | <0.001 | 0.814 (0.789 - 0.839) | 0.269 | 0.985 (0.960 - 1.012) | <0.001 | 0.912 (0.893 - 0.931) |
| Micropolitan areas | <0.001 | 0.600 (0.579 - 0.621) | <0.001 | 0.320 (0.281 - 0.365) | <0.001 | 0.398 (0.352 - 0.450) | <0.001 | 0.517 (0.487 - 0.550) |
| Not metropolitan or | <0.001 | 0.525 (0.490 - 0.562) | <0.001 | 0.238 (0.186 - 0.303) | <0.001 | 0.195 (0.143 - 0.267) | <0.001 | 0.465 (0.410 - 0.528) |
| micropolitan (non-urban residual) |  |  |  |  |  |  |  |  |
| **Hospital teaching status**<br>(Reference: Non-metropolitan hospital) |  |  |  |  |  |  |  |  |
| Metropolitan non-teaching hospital | <0.001 | 1.451 (1.402 - 1.502) | <0.001 | 1.822 (1.596 - 2.079) | <0.001 | 1.994 (1.759 - 2.262) | <0.001 | 1.349 (1.267 - 1.436) |
| Metropolitan teaching hospital | <0.001 | 1.699 (1.646 - 1.753) | <0.001 | 3.254 (2.900 - 3.651) | <0.001 | 2.905 (2.591 - 3.257) | <0.001 | 1.997 (1.889 - 2.111) |

Under the next category including ages 1-4 yrs old, the predominant causes for comorbidities associated with admission were cancer [6.217 (5.989 - 6.454), p<0.001], transplant recipients [1.942 (1.789 - 2.108), p<0.001], hepatitis[1.770 (1.096 - 2.859), p<0.001].Children admitted with causes such as asthma/bronchitis, pneumonia, diabetes and dehydration showed lesser chances of readmission (Figures 1,2,3 and 4).

**Figure 1.**
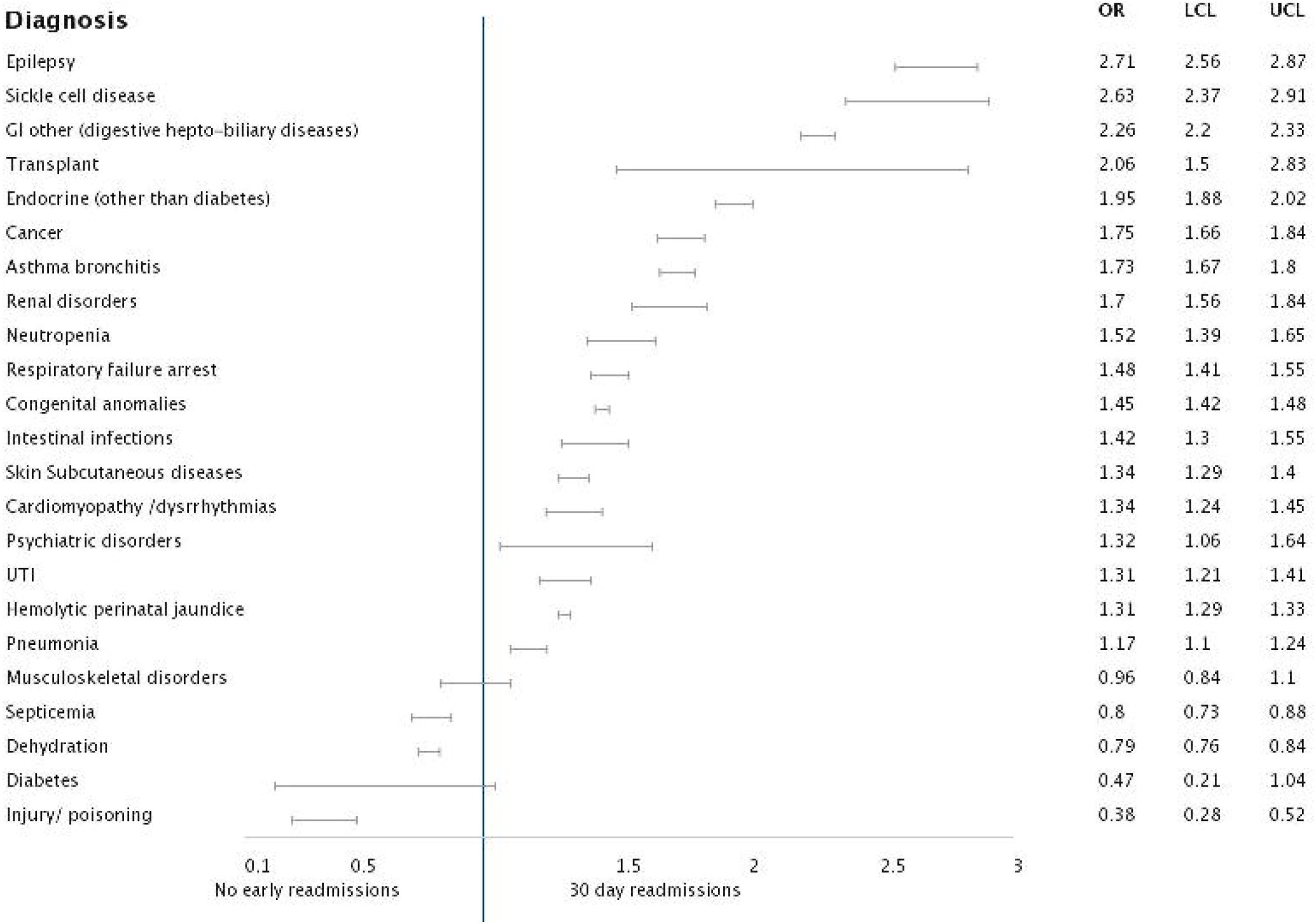
Forrest plot analysis of pertinent diagnosis associated with 30 days readmission: Age group <1years;

**Figure 2.**
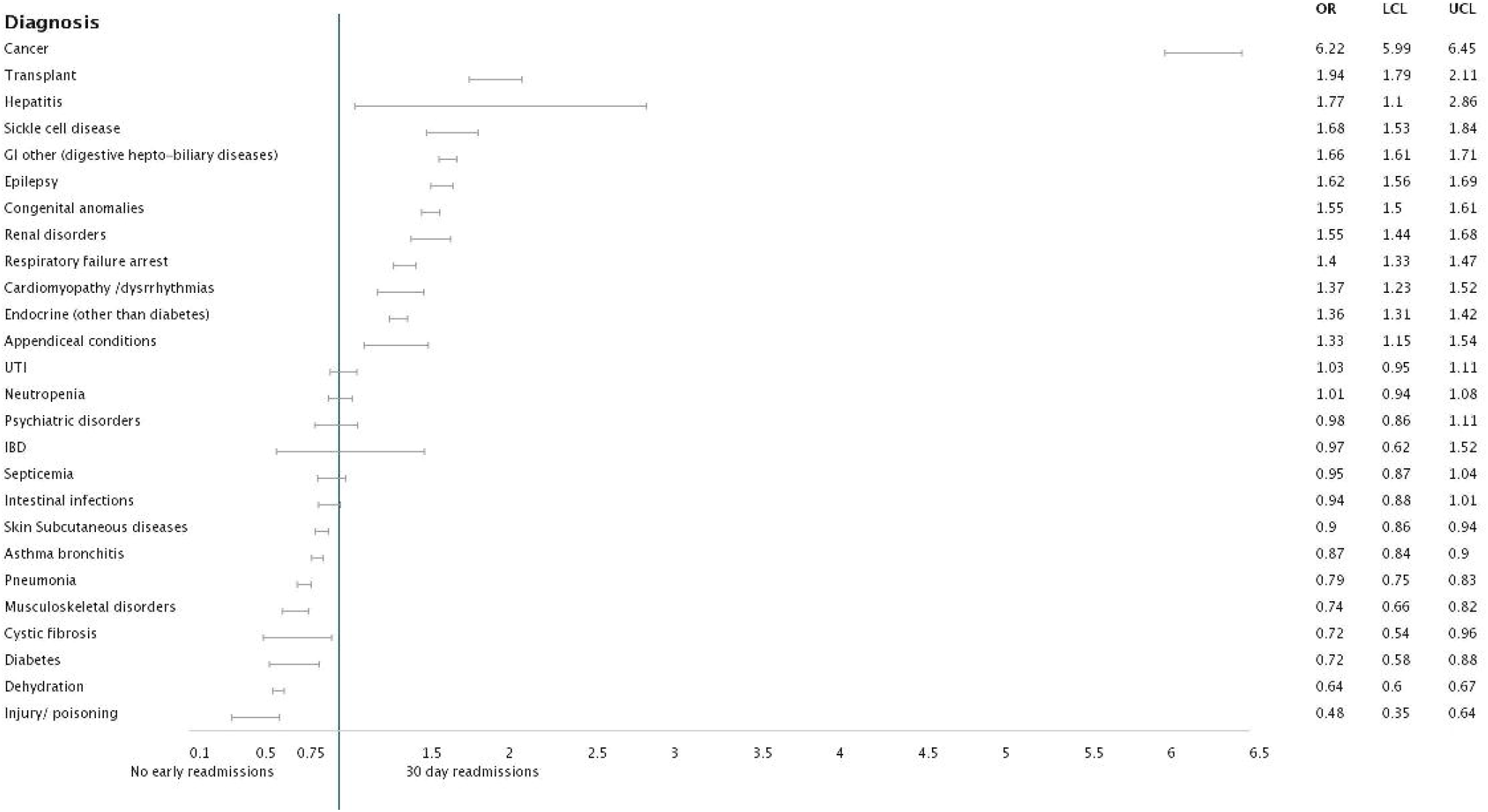
Forrest plot analysis of pertinent diagnosis associated with 30 days readmission: Age group 1-4 years;

**Figure 3.**
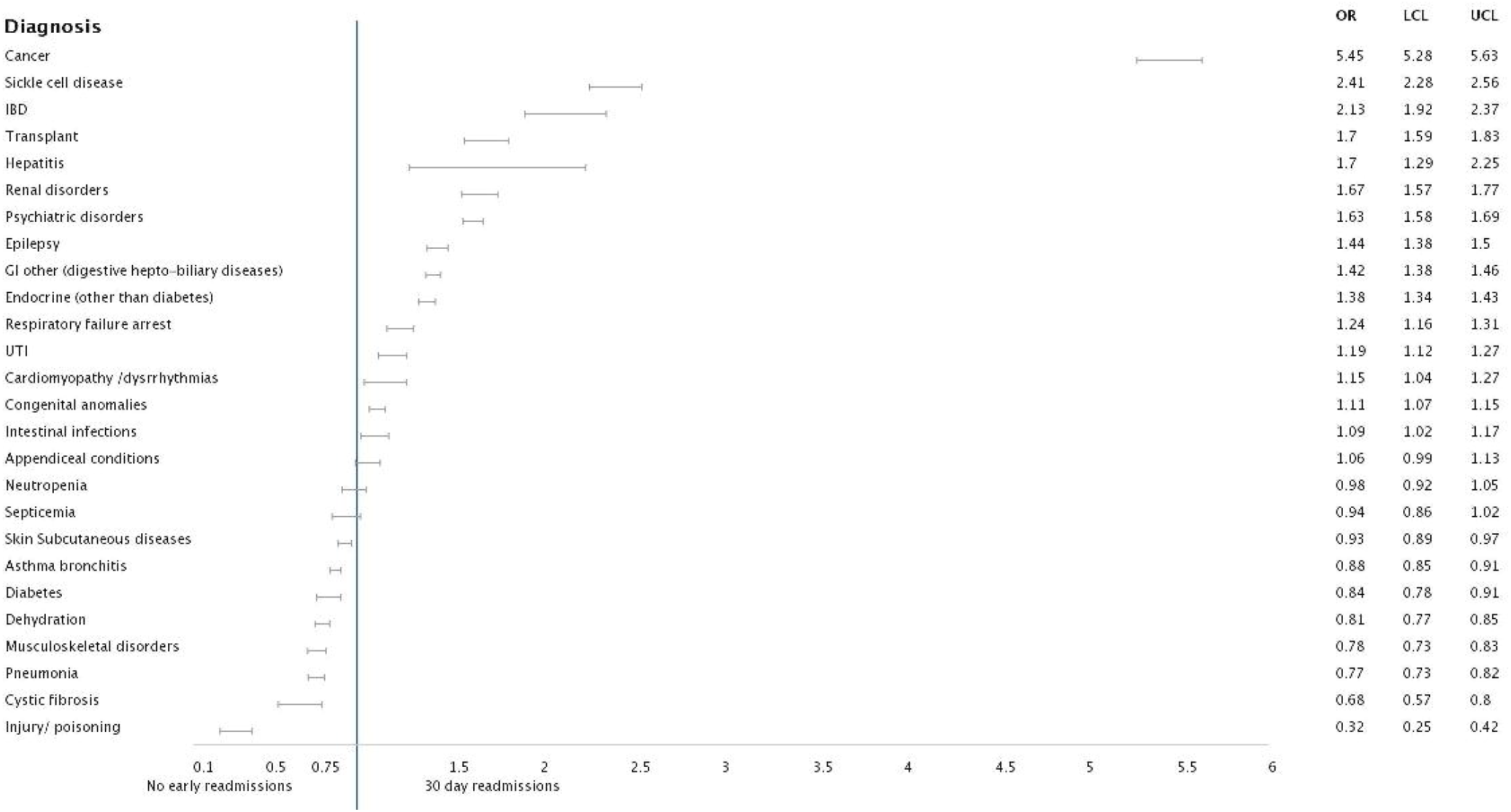
Forrest plot analysis of pertinent diagnosis associated with 30 days readmission: Age group 5-12 years;

**Figure 4.**
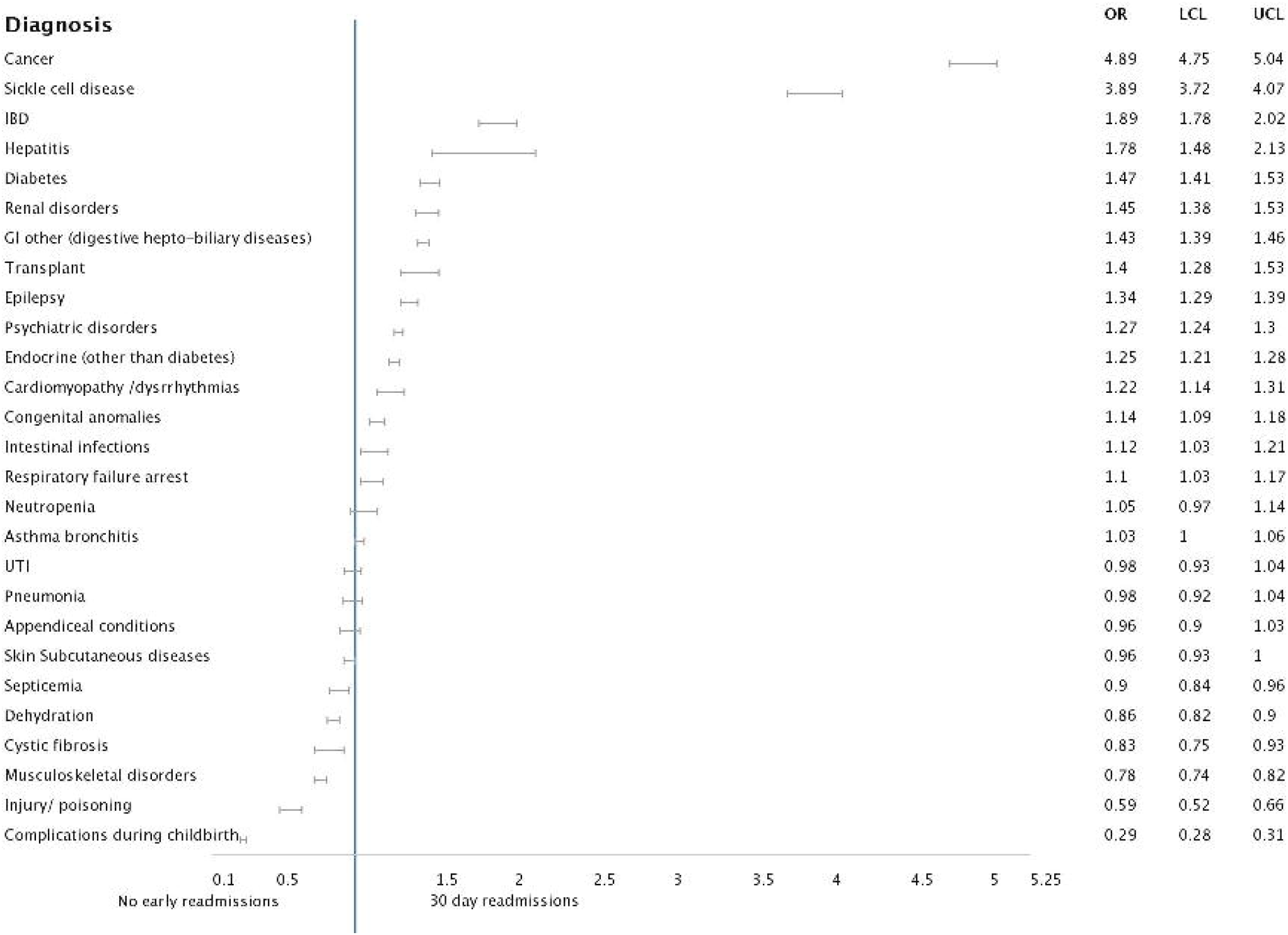
Forrest plot analysis of pertinent diagnosis associated with 30 days readmission: Age group 13 years and above

In the third category of ages between 5-12 years, the highest odds of occurrence of readmission were with comorbidities namely cancer [5.453 (5.278 - 5.634, p<0.001], sickle cell diseases [2.413 (2.275 - 2.560), p<0.001] and inflammatory bowel diseases (IBD) [2.131 (1.920 - 2.366), p<0.001]. There were a multitude of admission comorbidities with odds against readmission for this age-group, top causes of which included pneumonia, cystic fibrosis and musculoskeletal injuries.

For adolescent ages, the highest associations with readmission were under cancer [4.892 (4.745 - 5.043), p<0.001], sickle cell disease [3.890 (3.721 - 4.066), p<0.001], renal diseases [1.451 (1.381 - 1.525), p<0.001] and IBD[1.893 (1.778 - 2.017), p<0.001]. Another important association was psychiatric illnesses [1.272 (1.244 - 1.300), p<0.001] which had not been seen in the previous groups. The patients who showed a negative prediction for readmission were composed of groups with co-morbidities namely cystic fibrosis, musculoskeletal causes, skin diseases, dehydration and injuries/ poisonings.

## DISCUSSION

In this nationwide represented database for the pediatric population, we have the opportunity to analyze a large sample size which gives us a more reliable outcome. To the best of our knowledge this is a first pediatric, age defined (including infants), all cause, allpayer review of national data from the Healthcare Utilization Project, Nationwide Readmission Database using (around 5.5 million inpatient encounters with ICD-10 data.

Using nationally representative data for US rehospitalizations, the biggest trend we demonstrated through this study was increasing rates of unplanned readmissions with cancer as a comorbidity across all age groups. This corroborates with previous studies which has time and again shown cancer and transplant recipients to be a higher association for readmissions. [15] The total rate of readmission is 3.2% for all ages inclusive of ages less than 1. For those older than 1 year of age, the rate of readmission is 6.7%. In comparison to previous years (2010-2016) this number has evolved from 6.26 to 7.02 % to 6.7% in our study, hence there is some improvement in health care interventions and strategies on the national scale as seen by the improvement in rates of readmission. [16] Another independent study conducting readmission rate assessment for 72 free standing pediatric hospitals in 2013 showed a rate of 6.2% which might be attributed to the fact that the hospitals may not have had sicker patients at the time. [17]

While the causes are individually different under different age categories, there were a few patterns noted in our study. While chronic complex ailments such as sickle cell diseases and cancer have significant associated comorbidities which continue to show an increasing rate for ages above 1. Cohen et al and Sehgal et al had also demonstrated similar trends through their studies. [18, 19] Besides the illness severity which may be a valid cause, there is a continuous need for improvement in patient compliance and education for these groups of children which might help in improving readmission rates. [20] The prevalence of children with chronic conditions is increasing because of improved survival in the neonatal period and medical advances in care and technology ultimately leading to additional medical needs and inpatient stays in future. [21] By making comprehensive discharge protocols including provision of discharge medications in hand at the time of hospital discharge, dedicated education at the time of discharge and close follow up, unplanned readmissions can be potentially reduced. [22]

Bardach et al reported in 2013 that the pediatric condition specific readmission rates across various hospitals may not be a useful measure for comparison of performances since certain institutional places will always have a higher rate due to it being a bigger center for admission for especially chronic medical ailments. [23] They also suggested that few hospitals that care for children are identified more or less popular for revisits, even for common pediatric diagnoses, likely due to low hospital volumes. Bucholz et al previously described trends for readmission rates from 2010-2016.15 They determined that, although the total number of hospital admissions declined over these years, however the readmission rates were found to have increased. They also found that the higher readmission rates were associated with chronic diseases, which has been reiterated through our study with most contemporary data. Recently they also identified the impact of varying patient insurances over the rates of readmission. It was seen that the readmission rates for Medicaid- and privately insured pediatric patients declined slightly from 2010 to 2017 but continued to remain higher relatively and reiterated in our study amongst the Medicaid beneficiaries indicating the role of insurance affecting the services and need for rehospitalizations. [24]

It has been noted that in ages 1-4 years dehydration along with asthma/bronchitis which are one of the most common diagnoses in the pediatric population across the United States, are negative predictors of unplanned readmission. This might be possible due to adequate close follow up and use of preventive measures like Asthma Action plans, good asthma scoring by more and more institution can lead to reduction in readmissions. [25] Asthma by itself has however been shown to have higher rates of readmission but that may just be reflecting the negative predictor for this age group alone as illustrated from our study. [26] Psychiatric illnesses including anxiety disorders, depression and suicidal ideation tend to come up as one of the forthcoming causes in the older age groups which leads to higher rehospitalizations. This is another important focus for all future approaches and plans of action. Sehgal et al recently demonstrated urinary tract infections to be negative predictors of readmission for this age group which might also be attributed to close outpatient follow up which has been illustrated with our study. [19] Richards et al demonstrated the importance of the internet as a means of managing depressive symptoms through the CATCH-IT trial. They reported through their analysis that greater motivation for depression prevention and lower ratings of self-efficacy at baseline were associated with greater declines in depression symptoms. [27] Inflammatory bowel diseases are also a significant burden for this age group and amongst patients of Crohn’s disease, psychiatric causes are a frequent cause for readmission. [28]

Among infants, factors like hyperbilirubinemia and bronchiolitis are associated with increased unplanned readmissions. It may be interesting to consider sub-threshold phototherapy for neonates during birth hospitalizations as a measure which has been previously studied which might contribute to its overuse. [29] Readmission rate of bronchiolitis in previous studies have ranged from 1-4%, which when compared to overall pediatric readmissions appears to be on the lower spectrum. [30] The higher rates were also seen for reflux, milk protein allergy and other Gastrointestinal disorders in this group which is an important causal association and has been less studied due to lesser research for this age group.

There are certain factors that will always continue to affect outcomes but may not be accounted for such as parental perception. Multiple times it might be considered that a child is fit for discharge but a parent may feel that they are not healthy enough which is an important consideration for the pediatric population. [31] On the other hand there are certain factors which may feel that they may influence readmission but they actually do not affect largely. Krumholz et al also determined from their study that the rate of readmission is not related to increased mortality rates, by which we can safely say that early triaging and rapid management and interventions may not play a role in readmission rate. [32]

Moving forward, we need further investigations to understand the reasons for variation in readmission rates across children’s hospitals. There may be subtle variations in the form of care during the index hospitalization as well as post discharge care which could be a contributing factor. The bigger hospitals catering to a larger population have historically had higher rates of unplanned readmissions likely because they are more widely available and accessible. [33] Multiple variables at the patient, family and community level may influence readmission risk and thus require investigation. Use of artificial intelligence should be employed for predicting readmission rates for better validation and accuracy as demonstrated by Amritphale et al in their recent study. [34] This is one of the leading algorithms for research and is causing scientists to rethink how we integrate information and analyze data.

Furthermore, work is needed to determine how to measure the most relevant variables accurately and reliably using available data sources. Factors such as access to primary care for follow up visits or community factors such as parental ability to avail paid leaves may also play a role. In addition, there can be unpredictable disease progression which may govern increased length of stay and may affect readmission rates as well. [35]

NRD is in a format of annualized data where each patient could have a maximum follow-up duration of only 1 year and the same patient cannot be followed over the years even if data from multiple years is combined. By design, NRD does not allow determination of regional variations within the dataset. This study does not give a glimpse into patients with various causes as an outpatient procedure and represents only hospital admissions. As with any observational data, the results do not suggest causal relationship as there can be other unmeasured confounders. NRD data does not include the ‘observation stays’ in our analysis that might alter the total number of readmissions but would be unlikely to change the overall trend in readmission rates after inpatient admissions. Lastly, the NRD database does not provide pharmacological data that may impact readmissions. Also being an administrative database there is always a possibility that the ICD-10 codes may or may not be completely accurate.

## CONCLUSION

Our analysis shows the overall readmission rates are 3.2 % but for those older than 1 year of age, the rates were higher (6.7%). The chronic complex medical conditions, cancer and sickle cell disease account for greater readmission rate across all age groups along with psychiatric disorders and inflammatory bowel diseases being an important association for older age groups. This study will pave the way for more comprehensive strategies to help identify patients at high-risk of unplanned readmission for targeted interventions. This will also help control and increase savings to the wider health care economy with total charges for unplanned readmissions mounting to over $70 billion.

## Data Availability

All data produced in the present work are contained in the manuscript.

